# Non-Invasive Prenatal Diagnosis of Single Gene Disorders with enhanced Relative Haplotype Dosage Analysis for diagnostic implementation

**DOI:** 10.1101/2023.01.13.23284512

**Authors:** Mathilde Pacault, Camille Verebi, Magali Champion, Lucie Orhant, Alexandre Perrier, Emmanuelle Girodon, France Leturcq, Dominique Vidaud, Claude Férec, Thierry Bienvenu, Romain Daveau, Juliette Nectoux

## Abstract

Non-invasive prenatal diagnosis for single-gene disorders (SGD-NIPD) has been widely adopted by patients, but is mostly limited to the exclusion of paternal or de novo mutations. Indeed, it is still difficult to infer the inheritance of maternal allele from cell free DNA (cfDNA) analysis. Based on the study of maternal haplotypes imbalance in cfDNA, relative haplotype dosage (RHDO) was developed to address this challenge. Although RHDO has proven to be reliable, robust control of statistical error and explicit delineation of critical parameters for assessing the quality of analysis have not been fully considered yet.

Here we propose a universal and adaptable enhanced-RHDO procedure (eRHDO) through an automated bioinformatics pipeline with a didactical visualization of results that aims to be applied for any SGD-NIPD in routine care. A training cohort of 43 families carriers for *CFTR, NF1, DMD, or F8* mutations allowed the characterization and optimal setting of several adjustable data variables, such as minimal sequencing depth and type 1 and type 2 statistical errors, as well as the quality assessment for intermediate steps and final result through block score and concordance score. Validation was successfully carried out on 56 pregnancies of the test cohort. Finally, computer simulations were used to estimate the effect of fetal-fraction, sequencing depth and number of informative SNPs on the quality of results. Our workflow proved to be robust, as we obtained 94.9% conclusive and correctly inferred fetal genotypes, without any false negative or false positive result.

By standardizing data generation and analysis, we fully describe a turnkey protocol for laboratories wishing to offer eRHDO-based non-invasive prenatal diagnosis for single-gene disorders as an alternative to conventional prenatal diagnosis.

## Introduction

The identification of fetal DNA in maternal blood by Dennis Lo in 1997 has dramatically changed the landscape of prenatal diagnosis (PND).^1^ The major impact to date is probably the widespread use of cell-free DNA (cfDNA) testing for frequent fetal aneuploidies and, more recently, subchromosomal abnormalities,^2–5^ which was largely adopted by clinicians and pregnant women,^6^ and drastically reduced invasive sampling rate for this indication over a few years.^7,8^ Detection of fetal-specific sequences that are absent or different from maternal genome is also routinely performed, in the context of fetal sex determination,^9,10^ fetal RhD genotyping^11,12^ and exclusion of paternally-inherited or *de novo* mutations for fetuses at risk of single-gene disorders (SGD).^13–15^ However, determination of fetal status towards maternal pathogenic variants remains more challenging, as haploidentical maternal and fetal sequences cannot be easily distinguished.

Whereas detection of paternally inherited or *de novo* variants relies on a qualitative presence/absence approach, most workflows described to date for maternal inheritance determination rely on the fine quantification of each maternal haplotype, called relative haplotype dosage analysis (RHDO), initially described by Lo et al.^16^ In this approach, based on Massively Parallel Sequencing (MPS), parental, proband and cfDNA are sequenced in parallel at multiple polymorphic positions along the locus of interest. The two haplotypes of each parent are phased by comparison with the proband’s genotype. Paternal inheritance is then determined based on the qualitative detection of polymorphisms where the two paternal alleles can be distinguished, in other words where the father is heterozygous and the mother is homozygous, and both paternal alleles have been identified as linked to the at-risk or the normal haplotype. On the contrary, maternal inheritance is based on the calculation of an allelic ratio at positions where the two maternal alleles can be distinguished, that is to say where the mother is heterozygous and the father is homozygous. The detection of a haplotypic imbalance would reflect the fetal contribution, from which we may deduce the maternal haplotype that was transmitted to the fetus, normal or at-risk. By relying on indirect diagnosis, RHDO theoretically allows diagnosis for every family, regardless of the inheritance pattern or the type of molecular anomaly and has been successfully applied to several SGD.^17–21^ However, to our knowledge, clinical implementation in public health service laboratories around the world remains sparse, probably because quality controls and decisional thresholds remain complex to define.^22,23^

In this report, we describe our RHDO-based NIPD workflow, that we call *enhanced*-RHDO (eRHDO), where we explain in detail our informatics and statistical analyses with precise risks control and present a straightforward and comprehensive result visualization. Using computer simulations, we discuss the impact of the different biological and analytical parameters on the quality of the result, and propose thresholds and objective quality scores for easy implementation in a diagnosis laboratory.

## Materials and Methods

### Participants and sample processing

Pregnant women who are known carriers of cystic fibrosis (*CFTR* gene), neurofibromatosis type 1 (*NF1* gene), Duchenne/Becker muscular dystrophy (*DMD* gene) or hemophilia (*F8* and *F9* genes) were recruited nationwide through the DANNI and NID studies, which were ethically approved by the French “Comité Consultatif sur le Traitement de l’Information en matière de Recherche dans le domaine de la Santé” (ref. 13.386) and “Comité de Protection des Personnes” (ref. 2014-janvier-13465 and 29BRC18.0055). We separated our cohort in two groups. The first group, called “training cohort”, included families for whom only parental, fetus and cfDNA were available, meaning parental haplotypes were determined using the current pregnancy’s fetus information. Blood samples from women in this group were initially used to validate the efficiency, accuracy and multiplexing capacity of our method. When genomic DNA (gDNA) from a first child or close relative was available, pregnant women were included in the group we called “test cohort”. For these patients, we used the proband’s gDNA to identify each parental haplotypes, allowing these families to be tested in a real-life-like setting.

The DNA samples needed for each family included the cfDNA extracted from maternal plasma, the maternal and paternal gDNA extracted from leukocytes, the proband gDNA when available and the fetal gDNA from invasive sampling to confirm the NIPD result (Figure 1). All samples from the same family were processed simultaneously and pooled together prior to targeted capture enrichment and MPS. Details on DNA extraction and sample processing for MPS libraries preparations are reported in the supporting information. After targeted MPS, sequencing data from each DNA sample were used to perform eRHDO analysis and determine fetal inheritance at the locus of interest. The outcomes were set as haplotype A, B, C or D for the training cohort, as the aim was to refine analysis thresholds and determine whether the fetal haplotype was correctly inferred, regardless of its risk of SGD. For the test cohort, we studied only the gene involved in the familial SGD, and outcomes were set as “HapI” or “HapII” for maternal at-risk or non-at-risk haplotype, respectively; and “HapIII” or “HapIV” for paternal at-risk or non-at-risk haplotype, respectively.

**Figure 1.**
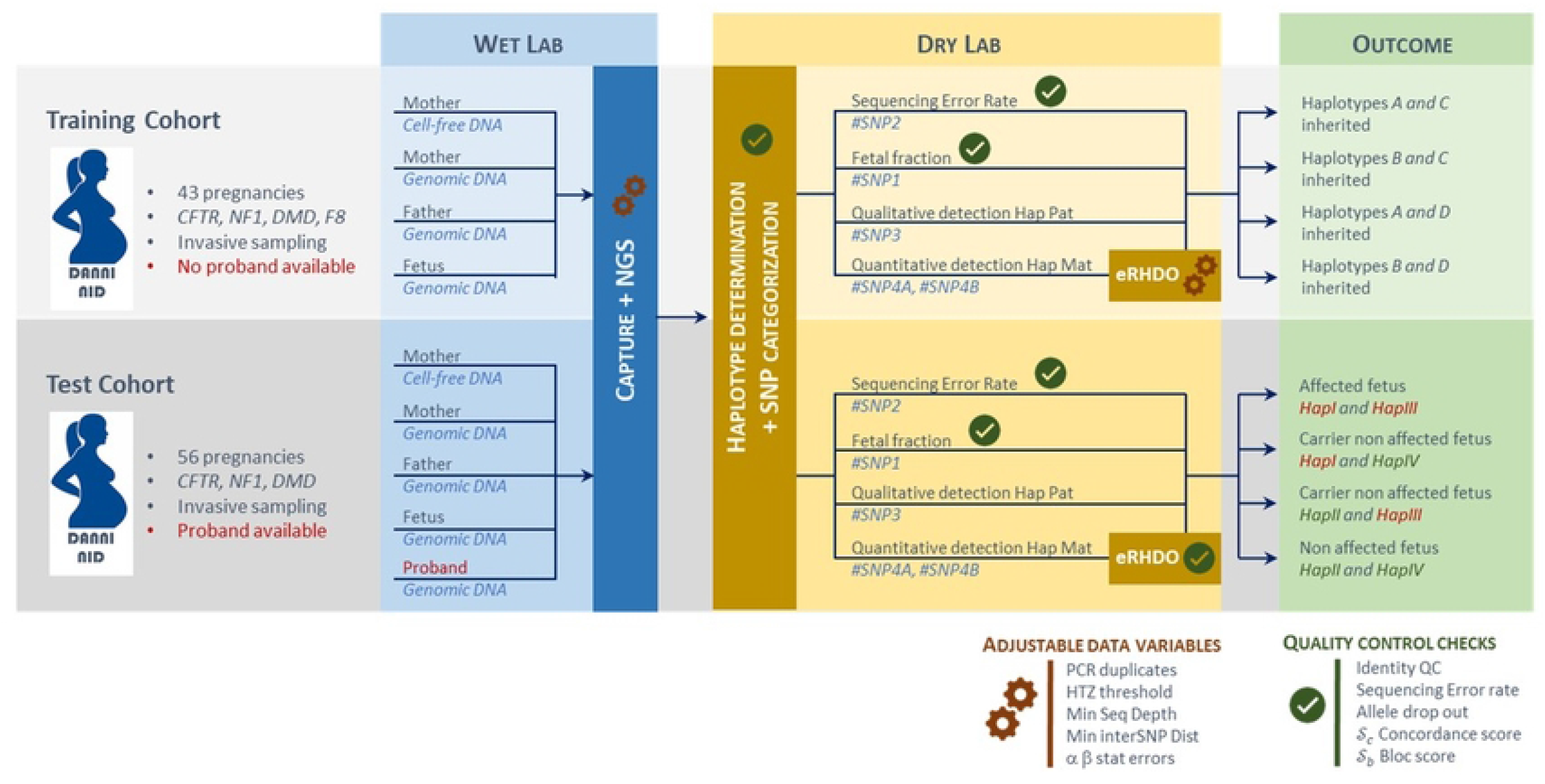
Workflow and processing steps of samples obtained from training and test cohorts. Training cohort involved couples at risk for SGD who were offered invasive prenatal testing and for whom parental haplotypes were determined using the current pregnancy’s fetus genomic DNA. Test cohort involved pregnant women at risk for SGD who were offered invasive prenatal testing and for whom a proband’s genomic DNA was available for haplotype reconstruction. Study of training cohort allowed to evaluate and set up the adjustable data variables. Then, the best combination of variables was used for test cohort exploration. Finally, the diagnostic performance of our SGD-NIPD workflow with eRHDO was evaluated and compared to gold standard as defined by fetal status obtained through invasive sampling. Quality controls checks are described to minimize mistakes and to help with technical validation and biological interpretation of the result.

#### SGD-NIPD Workflow: processing steps and terminology

After sequencing, a family-specific pile-up was generated, counting the number of reads for each base, from parental at-risk and non-at-risk haplotypes, at every SNP targeted in our capture panel. The two maternal haplotypes are HapI and HapII, HapI being the maternal at-risk haplotype and HapII the maternal non-at-risk haplotype; while the two paternal haplotypes are HapIII and HapIV, HapIII being the paternal at-risk haplotype and HapIV the paternal non-at-risk haplotype.

Each SNP of the pile-up was categorized based on parental inheritance, as initially described by Lo et al.^16^ Briefly, SNP1 are defined as those for which the father and mother are both homozygous for different alleles (mother AA and father BB). SNP2 are those in which the father and mother are both homozygous for the same allele (mother AA and father AA). SNP3 are those for which the father is heterozygous and the mother is homozygous (mother AA and father AB). Finally, SNP4 are those for which the father is homozygous and the mother is heterozygous (mother AB and father AA). SNP4 can be subclassified as SNP4a and SNP4b. SNP4a are those for which the paternal allele is the same as the maternal allele from HapI, while SNP4b are those for which the paternal allele is the same as the maternal allele from HapII. The other genomic positions of the pileup are categorized as WARNING_1 or WARNING_2 (Tables S1 and S2).

SNP from each category were then analyzed in maternal plasma through SGD-NIPD investigation to provide simultaneously i/ sequencing error rate determination (SNP2 analysis), ii/ fetal fraction evaluation (SNP1 analysis or uncategorized SNP analysis), iii/ qualitative paternal haplotype transmission determination (SNP3 analysis) and iv/ quantitative maternal haplotype transmission determination (SPRT-based eRHDO SNP4 analysis). After technical validation of the result, biological interpretation and medical conclusion can be conducted.

The SGD-NIPD workflow is reported in Figure 1.

### Data analysis

Using simulated cfDNA sequencing data and data from the training cohort, we defined adjustable data variables, namely consideration of PCR duplicates, thresholds used to discriminate a heterozygous position in gDNA and minimal sequencing depth for calling SNP in gDNA (NDP) and cfDNA (PDP). Minimal interSNP distance for eRHDO analysis, as well as type I and type II error risks associated with SPRT haplotype block delimitation were defined as adjustable output data variables. Afterwards, we tested different combinations for input and output data variables and examined objectively their impact on the quality scores in order to select the best combination of input and output data variables for test cohort exploration. Finally, the diagnostic performance of our noninvasive test was settled from the test cohort analysis.

Details of the bioinformatics pipeline for sequencing data analysis are reported in Supplemental Methods. We generated a graphical visualization of the results that include preanalytical and analytical information, such as input data (inheritance mode, proband gender for X-linked disorders), fetal fraction and quality parameters (position of the parental variant, total number of SNPs called for each analysis and distance between tested SNPs). For eRHDO analysis, the repartition of SNP4a and SN4b along the locus, the mean count of SNPs per haplotype block, the haplotype block count, the concordance between forward- and reverse-defined blocks (*pc*), as well as the proportion of positions where the SPRT remained non-conclusive (*nc*) are specified. SNPs were arbitrarily colored in green for the wild-type linked-haplotype, in red for the mutant-linked haplotype, and in gray for non-conclusive results. This whole analysis pipeline was created and refined using our training cohort. Its reliability and robustness, as well as pertinence of the chosen thresholds were tried on our test cohort. In this research study, all results were compared to the fetal genotype obtained on invasive sampling.

### Quality parameters for eRHDO analysis

We implemented quality controls tags as well as two eRHDO-specific quality scores, that we named concordance score *𝒮*_*c*_ and block score *𝒮*_*b*_, with the aim of reflecting forward and reverse concordance, haplotype classification error rate as well as the ease to conclude in favor of the over-representation of one of the maternal haplotype,

The concordance score *S*_*c*_ is defined as the difference between the concordance proportion between forward and reverse analyses *pc* and the proportion of SNPs that remained non-conclusive *nc*. To strongly discriminate against misclassified haplotype blocks (low *pc*) and low-quality results (high *nc*), which would reflect a low quality result to be interpreted with caution, *S*_*c*_ is calculated as follows: *S*_*c*_ = − *log*_100_(1 − *pc*) −*nc*. A *S*_*c*_ closer to 1 reflects a more robust result. The block score *𝒮*_*b*_ is obtained by normalizing the averaged number of SNPs per haplotype block by the total number of SNPs tested on the locus, expressed in a logarithmic scale for more discrimination: 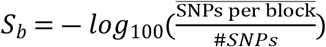. Shorter haplotype blocks means that fewer positions were tested before reaching statistical significance. In other words, a block score close to 1 reflects a bigger imbalance between both maternal alleles, therefore a higher confidence in the fetal haplotype that is called.

### Statistical analysis

All statistical analyses were performed using R. To measure the effect of the adjustable data variables on the results quality, we performed Wilcoxon tests and Friedman tests for variables with two (heterozygous position in gDNA) or more than two groups (minimal sequencing depth and minimal interSNP distance). In addition, we represented the associations using boxplots.

The effect of all quality-influencing factors (fetal fraction, number of SNPs and sequencing depth) and their combinations on the results quality was quantified using Sobol sensitivity indices, ranging between 0 (minor impact) and 100 (major impact). P-values for ANOVA tests were also computed to verify their significance. All p-values were adjusted for multiple comparisons using the Bonferroni correction.

## Results

### Patients information

The training cohort included 43 pregnancies, divided between 27 pregnancies at 25% risk of cystic fibrosis, 9 pregnancies at 50% risk of maternal transmission of neurofibromatosis type 1, 3 and 4 pregnancies involving a male fetus at 50% risk Duchenne/Becker Muscular Dystrophy and Hemophilia, respectively. Gestational age at sampling ranged from 6 to 29 weeks of gestation (mean, 12 weeks).

The test cohort included 56 pregnancies from 45 families, divided between 25 pregnancies at 25% risk of cystic fibrosis (paternal and maternal haplotypes transmission were non-invasively determined in an autosomal recessive pattern if both parents carried the same mutation, or with autosomal paternal + maternal dominant patterns if parents carried different mutations), 11 pregnancies at 50% risk of maternal transmission of neurofibromatosis type 1 (maternal haplotype transmission was non-invasively determined in an autosomal dominant pattern), and 20 pregnancies involving a male fetus at 50% risk of Duchenne/Becker Muscular Dystrophy (maternal haplotype transmission was non-invasively determined in an recessive X-linked pattern). Gestational age at sampling ranged from 6 to 17 weeks of gestation (mean, 11 weeks).

Patients’ information is summarized in Table S3.

### Optimal setting of the adjustable data variables

To refine our pipeline, the impact of several parameters on the quality of the result was tested using data from the training cohort.

#### Effect of PCR duplicates

As cfDNA has been largely shown to be non-randomly fragmented,^24,25^ two distinct fragments may be misinterpreted as PCR duplicates and eliminated on bioinformatics analyses, resulting in a loss of information and a risk of bias in the interpretation. To alleviate this risk, quality parameters of analyses were compared with or without PCR duplicates. As we obtained no difference in the fetal fraction, we settled on retaining the PCR duplicates removal in the final bioinformatics pipeline (data not shown).

#### Effect of the thresholds used to specify a heterozygous position in gDNA

We investigated two alternatives to define heterozygous SNPs in gDNA. In the extensive definition, heterozygous SNPs in gDNA are defined by an allelic frequency of the alternative allele (AF) comprised between 0.15 and 0.85, while homozygous positions in gDNA are defined by an AF <0.15 or >0.85. In contrast, the restrictive definition corresponds to heterozygous SNPs in gDNA defined by an AF comprised between 0.35 and 0.65 while homozygous positions’ AF in gDNA remain unchanged at <0.15 and >0.85. An extensive definition of a heterozygous position could retain more informative positions than the restrictive definition but is likely to introduce SNP categorization errors due to misspecification of SNP positions associated with biased allele frequency. We explored the effect of extensive versus restrictive definition of heterozygous SNPs in gDNA on the results by examining their impact on both scores (Figure S1). As expected, the use of the restrictive definition leads to a reduction in the number of SNP4 that can be used for the eRHDO analysis of the training cohort, which results in a lower *𝒮*_*b*_. On the other hand, this restrictive definition is associated with a significantly increased *𝒮*_*c*_, reflecting a marked improvement in the quality of the results (Table S4). Thus, we decided to opt for the restrictive definition.

#### Effect of the minimal sequencing depth

As a lower minimal sequencing depth threshold may retain information at more positions but is likely less precise in allelic ratio determination, we explored different minimal thresholds for sequencing depth for our gDNA and cfDNA analyses, denoted by gDNA sequencing depth (NDP) and cfDNA minimal sequencing depth (PDP), tested at 8x, 15x and 30x, and 15x, 30x, 45x and 60x, respectively. We examined objectively their impact on *𝒮*_*c*_ and *𝒮*_*b*_. At PDP fixed, we found no difference in our scores with higher NDP (Figure S2). Similarly, PDP variation did not impact the *𝒮*_*c*_ (Figure S3), indicating no impact on the concordance of haplotype blocks classification between forward and reverse orientations. However, we found significant differences in terms of *𝒮*_*b*_ (*p*-value for Friedman test of 4.8 × *e*^−24^ ; Table S5). *𝒮*_*b*_ value rises at low PDP, which reflects a better quality of analysis: the smaller the minimal sequencing depth for calling a SNP4 in cfDNA, the greater the number of SNP4 taken into consideration in the eRHDO analysis, with potentially a greater the number of blocks. Therefore, we fixed the minimal sequencing depth for calling SNP in cfDNA at 15x.

#### Effect of the threshold for minimal interSNP distance

Previous reports used a threshold for minimal interSNP distance of 200 base pair (bp) to minimize the risk of linkage between two SNPs and a resulting bias that would hinder allelic dosage analysis, but may also discard positions that would be informative.^18,19^ In order to evaluate this risk, we tested the impact of this threshold at 50 bp, 100 bp and 200 bp. The interSNP distance significantly impacted *𝒮*_*b*_, with a *p*-value for Friedman test of 3.6*e*^−46^, suggesting an increased distance between two SNPs might result in a loss of information by lowering the number of analyzed events (Figure S4 and Table S6). To maximize *𝒮*_*b*_, we settled for a minimal distance of 50 bp between two SNPs to be considered.

#### Effect of the statistical risks

When implementing the RHDO method, the statistical risks associated with the SPRT control the proportion of tolerated misclassification errors, thus having a major impact on both *𝒮*_*c*_ and *𝒮*_*b*_. Permissive risks, in other words risks closer to 1, make easier a conclusion of the SPRT test, which obviously leads to smaller haplotype blocks and better associated *𝒮*_*b*_ (Figure S5), but they have a variable impact on *𝒮*_*c*_ (Figure S6). For low values of the parameters fetal fraction, number of SNPs and depth of sequencing, obtaining conclusive results is rather challenging and higher values of the statistical risks, associated with smaller blocks, helps in that sense (Figure S7). However, in reasonable situations, the misclassification errors (Figure S8) induced by permissive statistical risks increases drastically. Therefore, we decided to set the two risks to 1/1000, with the aim of reducing at most the erroneous conclusions. On note that our pipeline allows to adjust each risk to other values if deemed necessary.

As a conclusion, the investigation of the training cohort enabled to test different combinations for input and output data variables, to objectively examine their impact on the quality scores and to pinpoint important quality control checkpoints. In fine, the optimal combination of input and output data variables for test cohort exploration corresponded to restrictive definition of heterozygous SNP, minimal NDP=8x, minimal PDP=15x, minimal interSNP distance=50bp, and type I and type II error risks associated with SPRT =1/1000.

### Quality assessment

Quality control points have been implemented at all stages of the workflow, to minimize the risk of error, but also to allow the users to assess the robustness and confidence they can have in their technical result.

#### Quality control tags associated with identity check

After at-risk and non-at-risk haplotype determination, SNPs are split into the previously described categories depending on the parental genotypes’ combination^16^. We adapted SNP categorization by considering the proband’s relationship to the couple and his status towards the familial SGD for subclassification, as well as their sex for X-linked transmission. Indeed, possible genotype combinations and decisional thresholds may differ whether the individual tested for parental haplotyping is a previous child or a relative of the couple. For quality purposes, we also included two “WARNING” categories, as consanguinity or undisclosed false paternity may hinder interpretation. Previously called SNP5, where both parents are heterozygous and the at-risk haplotypes cannot be inferred, were named “WARNING_2”, for “uninformative genotype combination”. Impossible genotypes combinations were called “WARNING_1”, for example parents “AA” and “BB” with a first child as a proband genotyped “AA”. This second category may be informative of a sample swap, a wrong proband relationship assignment, or low-quality sequencing (Tables S1 and S2).

#### Quality control tags associated with DNA capture and sequencing

Important allele drop-out (ADO) may result in erroneous determination of fetal fraction, lower confidence in paternal inheritance or hinder maternal haplotypes ratio determination. We therefore return an evaluation of the proportion of ADO using SNP1 (fetal genotype AB and maternal genotype AA), as the proportion of fetal specific B allele not detected in maternal plasma (AF of B allele in maternal plasma = 0). Although this parameter does not block subsequent SGD-NIPD analyses, a significant ADO should result in a cautious interpretation of the result.

Sequencing error rate for each family was estimated using SNP2 density plots, corresponding to positions where both parents are homozygous for the same allele: the fetus is expected to be homozygous as well and any other allele found at the locus can be considered a sequencing error. Here again, a significant sequencing error rate associated to one or more samples should result in a cautious interpretation of the result of the SGD-NIPD.

### Test cohort results

Patients’ information and eRHDO-based SGD-NIPD results are presented in Table S3.

Sequencing error rate was consistently low for all our families, ranging from 0.3% to 1.4% (mean, 0.68%, data not shown), consistent with the sequencing error profile of the platform.^26^ Although we chose not to set a fixed threshold for interpretation, an increased sequencing error rate should alert on the risk of low-quality result secondary to degraded DNA or a technical issue during DNA extraction or sample processing and sequencing.

Fetal fraction estimated using the distribution of all sequenced positions (minor allele frequency, MAF), as described in supplemental data, ranged from 2.9 to 19.3% (mean 9.3%). Three major discrepancies were found between MAF-based determination and SNP1-based determination, related to an abnormal distribution of the fetal-specific SNP1 allele frequency in maternal plasma caused by major ADO in these three samples (families 2_29, 2_33, 2_40).

An example of our result visualization is displayed in Figure 2.

**Figure 2.**
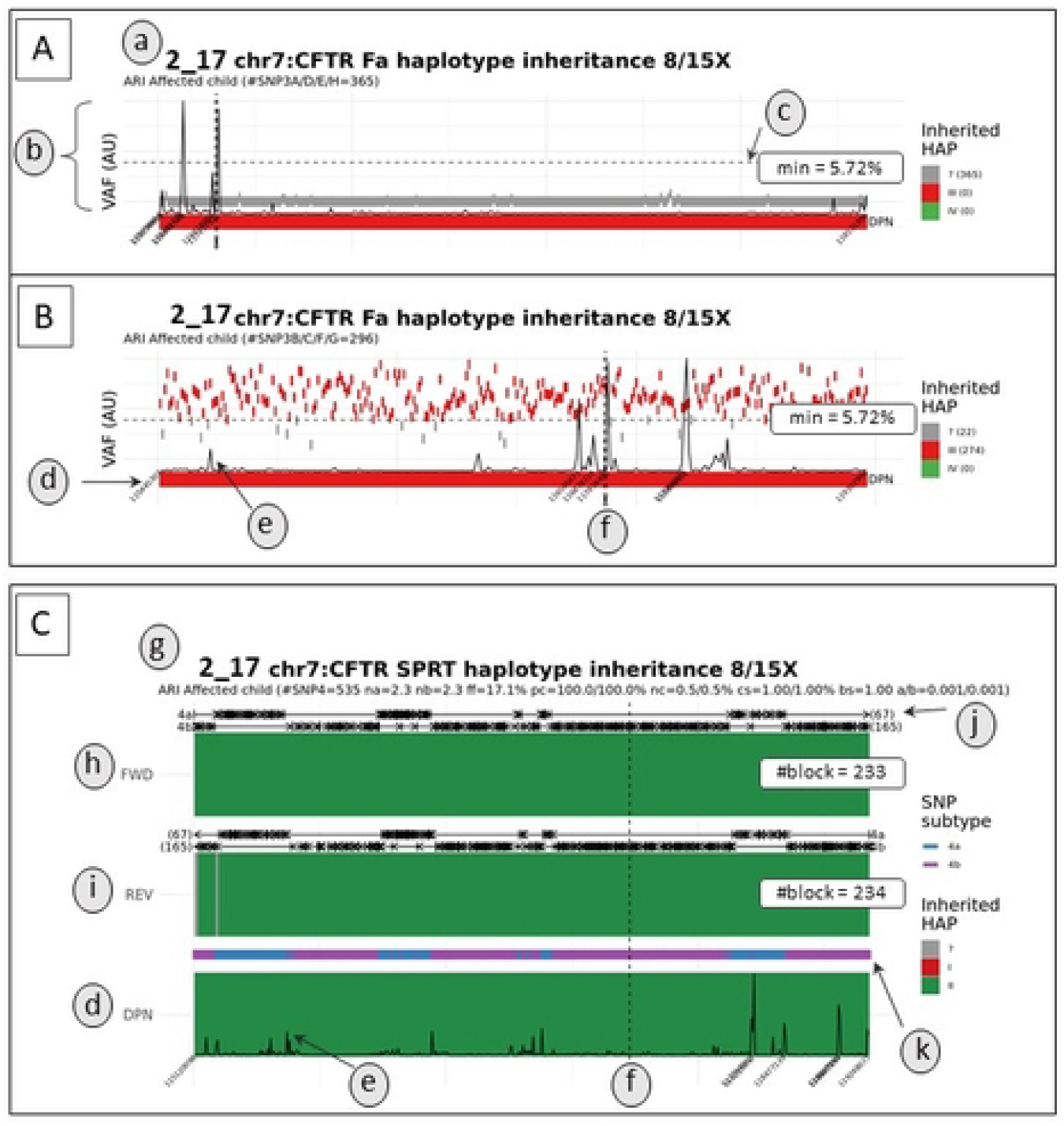
Example of a graphic result for autosomal inheritance in a family at risk of transmitting Cystic Fibrosis (family 2), subdivised in paternal (A, B) and maternal (C) inheritance. In this example, SNP3A/D/E/H correspond to SNP3 located on the “non at risk” paternal haplotype (A), whereas SNP3B/C/F/G correspond to SNP3 located on the “at risk” paternal haplotype (B). “Non at risk” and “at risk” haplotypes are represented in green and red, respectively. (a) For each result, input data are summarized as follow : family ID, tested locus and parental inheritance, gDNA/cfDNA minimal sequencing depth, transmission mode (ARI = autosomal recessive inheritance, ADI = autosomal dominant inheritance, RXI = recessive X-linked inheritance) and number of SNP3. (b) Qualitative detection of fetal-specific SNPs at an allelic frequency (VAF) higher than our threshold visualized in (c) (AU = arbitrary units). SNP3A/D/E/H from the “non at risk haplotype” were not detected in cfDNA (top), whereas the majority of SNP3B/C/F/G from the “at risk” haplotype where qualitatively detected above the background threshold (bottom). (d) Fetal genotype from sequencing of fetal gDNA obtained through invasive sampling (e) Representation of inter-SNP distance. Each peaks indicates a longer genomic distance between two consecutive tested positions. (f) Location of the parental mutation, symbolized by a vertical dashed line. (g) Input data and quality parameters of the SPRT analysis, namely number of SNP4, mean number of SNPs 4α or 4β per block (na and nb), concordance between conclusive haplotype blocks in forward and reverse orientation (pc) and proportion of non conclusive haplotype blocks (in grey color) among all blocks (nc), and type I and type II statistical error used in SPRT test (a/b). (h) and (i) SPRT analysis in forward and reverse orientation, respectively. (j) Visualization of haplotype blocks, divided in 4α (top) and 4β (bottom) analyses, with number of conclusive blocks. (k) Distribution of SNPs 4α (blue) and 4β (purple) along the genomic region.

Paternal inheritance was determined only for families at risk of recessive cystic fibrosis (CF). All results returned conclusive and concordant with the fetal genotype obtained on invasive sampling except one. In family 2_12, a recombination occurred close to the paternal variant location and the transmitted paternal haplotype at this position could not be inferred. Although the mother was a carrier of a different pathogenic variant, as this position was not targeted by our capture panel, the paternal variant could not be qualitatively detected by direct visualization of the paternal-specific allele on the bam sequence files.

Considering maternal inheritance, 59 analyses were performed on 56 samples, as one family (family 2_26) was included three times during three successive pregnancies in our study and the cfDNA of one given pregnancy was analyzed twice using either PND’s gDNA for parental haplotype inference. 56/59 samples returned conclusive (94.9%) and concordant with the fetal genotype obtained on invasive sampling. Among these, 43/59 results (72.9%) returned conclusive with high quality results. All these high quality results displayed a *𝒮*_*c*_ and a *𝒮*_*b*_ higher than 0.7 (Figure 3A). 13/59 (22%) returned conclusive and concordant with the fetal genotype but with slightly lower quality results. As suggested by our computer simulations, these low-quality results probably happened because of a small number of tested events, either because of low cfDNA sequencing depth, consistently lower than 100x (meaning a small number of events counted at each locus), or low fetal fraction (families 2_33 and 2_40) (meaning a small allelic imbalance to be detected). For these families, the block score *𝒮*_*b*_ returned between 0.55 and 0.7 (Figure 3B). Finally, 3 samples returned inconclusive (5.1%). For family 2_39, a recombination event occurred in the block adjacent to the position of the maternal variant, not allowing a confident haplotype inference at this position (Figure 3C). Family 2_12 had a low fetal fraction and a low overall cfDNA sequencing depth, so that not enough positions could be studied. Similarly, not enough SNP4 were included in family 2_14’s RHDO analysis as few SNP4 were identified in the *CFTR* locus and the sample returned with a relatively low fetal fraction (5.9%).

**Figure 3.**
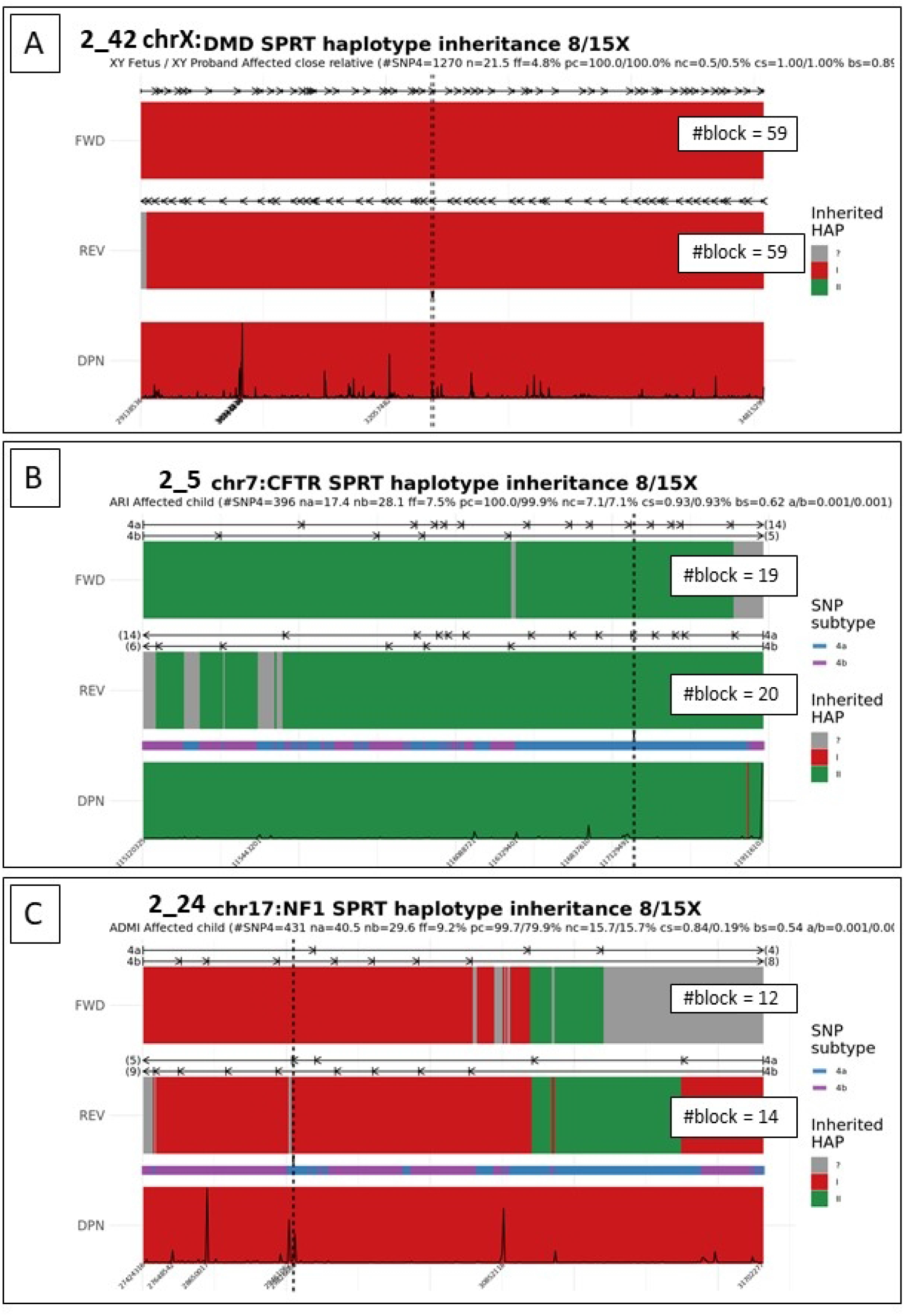
Examples of results with varying quality scores. (A) Family 2_42 at risk of transmitting Duchenne/Becker muscular dystrophy. A high number of tested SNP4 (n=1270) associated with a deep sequencing depth (mean=211X, data not shown) allowed determination of a great number of blocks (59 haplotype blocks in forward direction and 59 haplotype blocks in reverse direction) with high concordance in favor of the transmission of HapI (Sc=1.00), despite a relatively low fetal fraction (f=4.8%). Sb was evaluated at 0.89. (B) Family 2_5 at risk of transmitting Cystic Fibrosis. This family presented with a reasonable number of SNP4 (n=396) and fetal fraction (f=7.5%) but only a few haplotype blocks could be reconstructed (n=39 blocks in forward + reverse directions) with a relatively high number of SNP4 per block (na=17, nb=28.1), resulting in a lower block score (Sb=0.62). On note that the high concordance between forward and reverse analysis (Sc=0.93), as well as the position of the maternal variant along the CFTR locus (black dashed line) still allows to conclude in favor or the transmission of HapII with high confidence. (C) Family 2_24 at risk of maternal transmission of neurofibromatosis type I. Only 26 haplotype blocks could be defined with a high mean number of SNPs per block (na=40.5, nb=29,6), resulting in longer blocks and therefore a lower block score (Sb=0.54). Because 2/26 blocks were uncorrectly classified HapII instead of HapI, suggesting a SPRT error, the concordance score estimated at 0.84. Although the maternal variant is located in a concordant, HapI region, the low number of haplotype blocks, the haplotypic change at the 3’ end of the locus and the unclassified blocks at both extremities prevent us from concluding on the fetal status for NF1. In a diagnosis setting, analysis could be repeated on a subsequent sample. Sb : Bloc Score; Sc : Concordance score; na : mean number of SNP4a per haplotype bloc; nb : mean number of SNP4b per haplotype bloc; f : fetal fraction

We suggest that the use of thresholds determined by the user could help them when interpreting their result. In our hands, we observed that scores >0.7 were associated with high quality results, while scores <0.55 were associated with poor quality results.

### Computer simulation for quantification of the respective effect of quality-influencing factors

Thanks to both training and test cohorts’ study, we picked out biological and analytical critical factors that appear crucial for the quality of the analysis, namely fetal fraction, number of SNP4 available for eRHDO analysis, and cfDNA sequencing depth. To evaluate their respective relevance, we simulated sequencing data on which we tested the quality scores. In each simulation, one of the parameters was altered, the statistical risks being fixed to 1/1000.

As shown in Figure S9, these three parameters have a confirmed major influence on the quality of the results: the higher, the easier the conclusions of the SPRT tests, as well as the smaller and the more conclusive the blocks, the higher *𝒮*_*c*_ and *𝒮*_*b*_.

To quantify the effect of all these parameters on the result quality, we also performed sensitivity analysis through the computation of Sobol sensitivity indices. In brief, these indices measure how much the variance in the model output is due to each parameter to be fixed. A low sensitivity index (close to 0) reflects that variations of the associated parameter will lead to low variations in the results whereas a high sensitivity index (close to 100) implies huge changes. Sensitivity analysis also gives the opportunity to explore the interaction effects of parameters on the model. In addition to the computation of Sobol indices, we finally performed ANalysis Of VAriance (ANOVA) tests to identify the statistically significant effects of the parameters on both scores. Sobol indices of fetal fraction, number of SNPs, sequencing depth, combinations of them and the associated ANOVA *p*-values are provided in Table S7.

These further analyses confirm the major impact of the fetal fraction on both *𝒮*_*c*_ and *𝒮*_*b*_. The effect of the number of SNPs and the sequencing depth, while still being highly significant, is however less strong. Only interactions between parameters including the fetal fraction leads to significant variations of the *𝒮*_*c*_, which emphasizes again its importance. On the contrary, the *𝒮*_*b*_ is sensitive to simultaneous changes of all parameters.

## Discussion

Since the first description of RHDO, no practical recommendations or guidelines concerning data generation and analysis, fetal fraction evaluation, quality thresholds and interpretation have been proposed.^27^ In this report, we sought to develop a standardized workflow for clinical implementation of a reliable SGD-NIPD. To that effect, we worked on each step of the previously described RHDO and optimized this workflow on a training cohort, composed of families where the fetal gDNA was used to infer parental haplotypes. Then, we tried it on a test cohort, as close as possible to a clinical setting with various transmission modes and different types of molecular abnormalities as well as multiple proband and couple families’ schemes.

As we aim to provide the most direct computational analysis, we tried to review every situation that could be encountered during genetic counselling in our workflow construction. We detailed each SNP category by taking into account the proband’s relationship to the couple (previous child or close relative) as well as his status towards the parental variants (affected/carrier or unaffected). That way, haplotype phasing and decisional threshold calculations can be run automatically without the need for a bespoke adaptation of the informatics analysis pipeline for specific situations.

A key point of our approach is that our workflow has been designed in a way that users are able to adjust several input and output data variables at diverse steps of the process, according to their needs. The investigation of the training cohort as well as simulated data analysis enabled to test different values for these variables, to examine objectively their impact on the quality scores. Thereby, the removal or not of PCR duplicates, the extensive or restrictive thresholds used to specify a heterozygous position in gDNA, the minimum sequencing depth of cfDNA, the minimum interSNP distance, and the statistical risks associated with SPRT have been investigated and adjusted.

Quality control points have been implemented at all stages of the workflow, to minimize the risk of errors, but also to allow the users to assess the robustness and confidence they can have in their result. During SNP categorization, we added two “WARNING” categories for uninformative and impossible genotype combinations. The former may help expand the range of family members that may be tested, while the latter will be of use in identifying the potential causes of an unusual result. As an example, too many SNPs excluded from the analysis because of uninformative combinations may explain an inconclusive result on qualitatively correct experimental data that might reflect analysis from a consanguineous parental couple. An over-representation of impossible combinations may indicate an error in the input data for sequencing analysis, a sample swap or contamination during the technical steps.

Because ADO may hinder analysis, or even cause a false result, we added an evaluation of the mean ADO rate based on the SNP1 analysis in the output data. Finally, sequencing error rate density for each family was estimated using SNP2, corresponding to positions where both parents are homozygous for the same allele. Significant ADO or sequencing error rate should result in a cautious interpretation of the result.

The main quality-influencing factors are the fetal fraction, the number of SNP4 tested and the cfDNA sequencing depth, which impact the number of events tested. Computer simulations allowed identifying the fetal fraction as the most preponderant of them. As the number of SNP4 depends on the parental genotypes combination and fetal fraction is biologically determined in a given cfDNA sample, we recommend that sequencing conditions should be optimized in order to obtain sufficient cfDNA sequencing depth, higher that 100x in this study.

As fetal fraction needs to be evaluated precisely, we chose to use the minor allele frequency distribution of all the SNPs targeted in the capture panel rather than relying on the previous knowledge of their category, as it is done for SNP1-based evaluation. By increasing the number of events, we hope to obtain a more accurate estimate of the fetal fraction than the SNP1-based evaluation.

Paternal inheritance determination relies on the qualitative detection of fetal-specific SNPs that are present at a very low allelic fraction in maternal plasma. We introduced a minimal allelic fraction threshold for considering a SNP3 as significantly detected, depending on the sequencing error rate and the fetal fraction of the sample, to discriminate between background noise and SNPs that were indeed transmitted to the fetus. We also divided the analysis between “non-at-risk haplotype detected” and “at-risk haplotype detected” and require concordance between these two analyses.

Finally, we designed our output to be as simple and unambiguous as possible, while retaining as much information as possible. To that effect, we included a graphical visualization that summarizes the input data, reports the result with a two-color code, and includes several quality parameters to facilitate biological interpretation. In order to address the risk of misclassification due to an undetected recombination event, we represented the distance between two adjacent SNPs included in the analysis as a black curve in this final visualization, as the risk of recombination between two loci increases with genomic distance.^28,29^

Our workflow proved very robust, as we obtained 94.9% conclusive and correctly inferred fetal genotypes in our real-life-like cohort, and no false negative or false positive result was returned (56/59 cases conclusive and correctly inferred + 3/59 inconclusive cases). Rather, our quality controls proved stringent enough for low quality analyses to return inconclusive, mostly in the situation of an insufficient number of tested events, either because of a low fetal fraction, low sequencing depth or because of a small number of informative SNPs in the families. Our objective quality scores corroborated consistently our biological interpretation, being higher than 0.7 for high-quality analyses and lower than 0.55 for low-quality analyses. Nevertheless, we chose not to set a threshold for analysis, as we developed these quality scores as a support for interpretation rather than strict quality criteria. Indeed, they only refer to the quality of the haplotype identification in maternal plasma and do not consider the variant’s position along the target locus. They might therefore be close to 1 in a family for whom no SNPs 5’ or 3’ of the variant can be identified, a situation in which the maternal haplotype at the variant’s locus cannot be determined with confidence as a recombination event cannot be excluded. Likewise, if a recombination event is identified close to the parental variant position, the analysis may be of overall high quality but the interpretation of the SGD-NIPD should return inconclusive regardless of the quality scores.

This recombination risk is a well-described limitation of the MPS-based NIPD. Because of its reliance on an indirect approach without the possibility of the fetal sequences in the cfDNA, the occurrence of a recombination event close to the parental variant position may not allow to conclude.^23^ While this limitation cannot be entirely addressed, we hoped to assist interpretation by adding information on the parental variant position along the haplotype. In our test cohort, we observed a total of 7 recombination events, as expected from the known recombination frequencies in the genomic regions investigated. Five of them occurred at sufficient genomic distance of the parental variant and did not affect the SGD-NIPD result, but the other two hindered SGD-NIPD interpretation. On note, while a recombination event on the maternal allele close to the familial variant in the context of maternal autosomal dominant or X-linked transmission would not allow NIPD, if both parents carry different pathogenic variants in an autosomal recessive setting, paternal allele may be sought for directly in maternal cfDNA sequences in a qualitative manner.

Another major limitation of haplotype-based SGD-NIPD is the need for a proband’s genotype information for haplotype phasing. Caution must be made when choosing this proband. In some cases, one of the parents and their unaffected sibling may not share any haplotype. Similarly, when parents carry the same variant in an autosomal recessive disorder, the parental origin of the variant of an heterozygous first child will most likely be unknown, unless an indirect technique has been previously performed in these families. Parental haplotypes will be correctly phased but identification of the at-risk and non-at-risk haplotypes will not be possible. Although we anticipated most situations, this workflow cannot be offered to a family where a variant appeared *de novo*, or if no other family member is available for testing. Whereas *de novo* haplotyping methods have been reported in the context of NIPD,^30–35^ experimental approaches often require specific equipment and reagents, making them costly and inappropriate for implementation in a clinical setting. Computational approaches, while accurate and less expensive, may require specific bioinformatics skills and rely on population database, making them unreliable for rare or private variants phasing.^36,37^ An exception would be the Targeted Locus Amplification approach, described by Vermeulen et al, which does not depend on any specific equipment but may still require adaptations for implementation in laboratories unused to chromosome conformation capture techniques.^38^ Yet, experiences in preimplantation genetic testing showed that declining a referral because of the impossibility of analyzing a proband seldom happens.^39^

MPS-based techniques also require a careful study of the genomic environment of the tested gene before designing the enrichment panel. In our study, we tested hemophilia A-carriers in our training cohort, but had to exclude them from our test cohort, as our capture probes did not target enough SNPs at the 3’ extremity of the *F8* gene, which is localized at the telomeric extremity of the X chromosome. Specific adaptations may be needed for particular disorders, for example increasing the number of targeted SNPs in the locus of interest by lowering the minor allele frequency threshold used in the capture probes design, or increasing the size of the target region around the gene.

With the wide implementation of non-invasive prenatal testing for fetal aneuploidy, maternal incidental findings have been reported, especially maternal malignancy.^40,41^ To our knowledge, this situation has not been reported yet in the context of NIPD for SGD. The occurrence of an abnormal profile of our density plot of MAF for fetal fraction estimation may be suggestive of a maternal underlying disorder. To date, disclosure of results suggesting maternal malignancy remains controversial,^41,42^ but the possibility of maternal incidental findings during cfDNA analysis should be mentioned during pretest counseling.

As a conclusion, we report an easy-to-use workflow for SGD-NIPD. We adapted the previously published relative haplotype dosage analysis approach for a straightforward, easy to perform workflow. Although thresholds for statistical risks and quality parameters can be easily adjusted depending on the biological expertise, we propose an optimized value for each parameter. The accuracy and reliability of the whole process was then validated on a large real-life-like cohort. By testing multiple transmission modes, variant types and proband’s relationship, we hope to review a broad spectrum of situations frequently encountered in the PND context. This safe, non-invasive approach can be readily implemented as an accredited diagnosis service in a public health laboratory.

## Data Availability

All relevant data are within the manuscript and its Supporting Information files.

https://github.com/rdaveau/nipt_sprt_erhdo

## Acknowledgments

We are grateful to the patients, patients’ associations and the professional, geneticists, obstetricians, mid wives, nurses and laboratory technicians for supporting us throughout our developments. Thank you very much to Pr. Jean-Claude Kaplan and Dr. Christine Bellanne Chantelot for their wise and beneficial proofreading. The DANNI study received financial support from Agence de la Biomédecine, the association Vaincre la Mucoviscidose and the Association Française contre les Myopathies. The NID study received financial support from Agence de la Biomédecine and the association Gaétan Saleün.

## Author contributions

MP: conceptualization, methodology, validation, investigation, data curation, writing, visualization, funding acquisition; CV: conceptualization, methodology, validation, investigation, data curation, writing, visualization. MC: methodology, software, validation, resources, formal analysis, data curation, writing, visualization; LO: investigation; AP, EG, FL, DV, CF, TB: writing - review and editing; RD: methodology, software, validation, resources, data curation, validation, writing, visualization; JN: conceptualization, methodology, validation, resources, writing, visualization, visualization, supervision, project administration, funding acquisition.

## Data and code availability

Our NIPD pipeline is available on Github at https://github.com/rdaveau/nipt_sprt_erhdo.

